# New Cardiovascular Medications: Surveying Adoption Trends Among Cardiologists

**DOI:** 10.1101/2025.01.14.25320511

**Authors:** Leon Varjabedian, Abbas Mohammadi, Ashkan Mohammadi, Yasaman Vaseghi

## Abstract

**Introduction:** This study investigates factors influencing the adoption of new cardiovascular medications by private practice cardiologists in Las Vegas, focusing on economic, clinical, and informational barriers. Understanding these challenges is critical for improving medication uptake and optimizing patient care.

**Methods:** A cross-sectional survey was conducted between January 1 and February 1, 2024, targeting private practice cardiologists in Las Vegas. Out of 47 invited participants, 68% completed the questionnaire. The survey evaluated economic barriers, patient considerations, and preferred methods of learning about new treatments.

**Results:** The survey revealed that 54% of cardiologists adopt new medications within 2–6 months of approval, while fewer adopt them within 2 months or after 6–12 months. Economic barriers, particularly prior authorization requirements and high patient copays, were the most frequently reported challenges, surpassing concerns about clinical benefits or medication safety. Traditional conferences and professional society events emerged as the preferred sources of information for learning about new treatments, significantly outweighing reliance on digital platforms or pharmaceutical representatives.

**Discussion:** Economic factors, such as prior authorizations and copays, significantly delay the adoption of new cardiovascular medications. Cardiologists show a strong preference for formal educational resources, such as conferences and professional society events, over pharmaceutical outreach or digital platforms. Addressing these barriers requires streamlining administrative processes, reducing patient copays, and improving access to high-quality educational events. Collaboration between insurers, medical societies, and healthcare providers is essential to overcoming these obstacles and enhancing patient outcomes.

## Introduction

In the rapidly evolving field of cardiology, groundbreaking medications are reshaping clinical practice, supported by extensive clinical trials and FDA approvals ^1^. These advancements have targeted chronic conditions that significantly impact global healthcare costs, morbidity, and mortality. In the U.S., heart failure accounts for $39.2 to $60 billion in annual healthcare costs, and hyperlipidemia-related expenses reach $15.47 billion, with over half of American adults having high LDL levels (Fig 1) ^2,3^. Projections suggest heart failure’s economic burden could rise to $70 billion by 2030, with pharmaceutical costs for 2024 at $5 billion and undefined for 2030 ^4^. Hyperlipidemia’s projected costs for 2030 are about $11–$16 billion, considering a 2.5% growth rate (Fig 1) ^5^. These estimates, based on modeling and historical trends, underscore the urgent need for effective management strategies to reduce healthcare expenses and improve patient outcomes, highlighting the financial strain caused by heart failure and dyslipidemia in the healthcare system.

**Figure 1.**
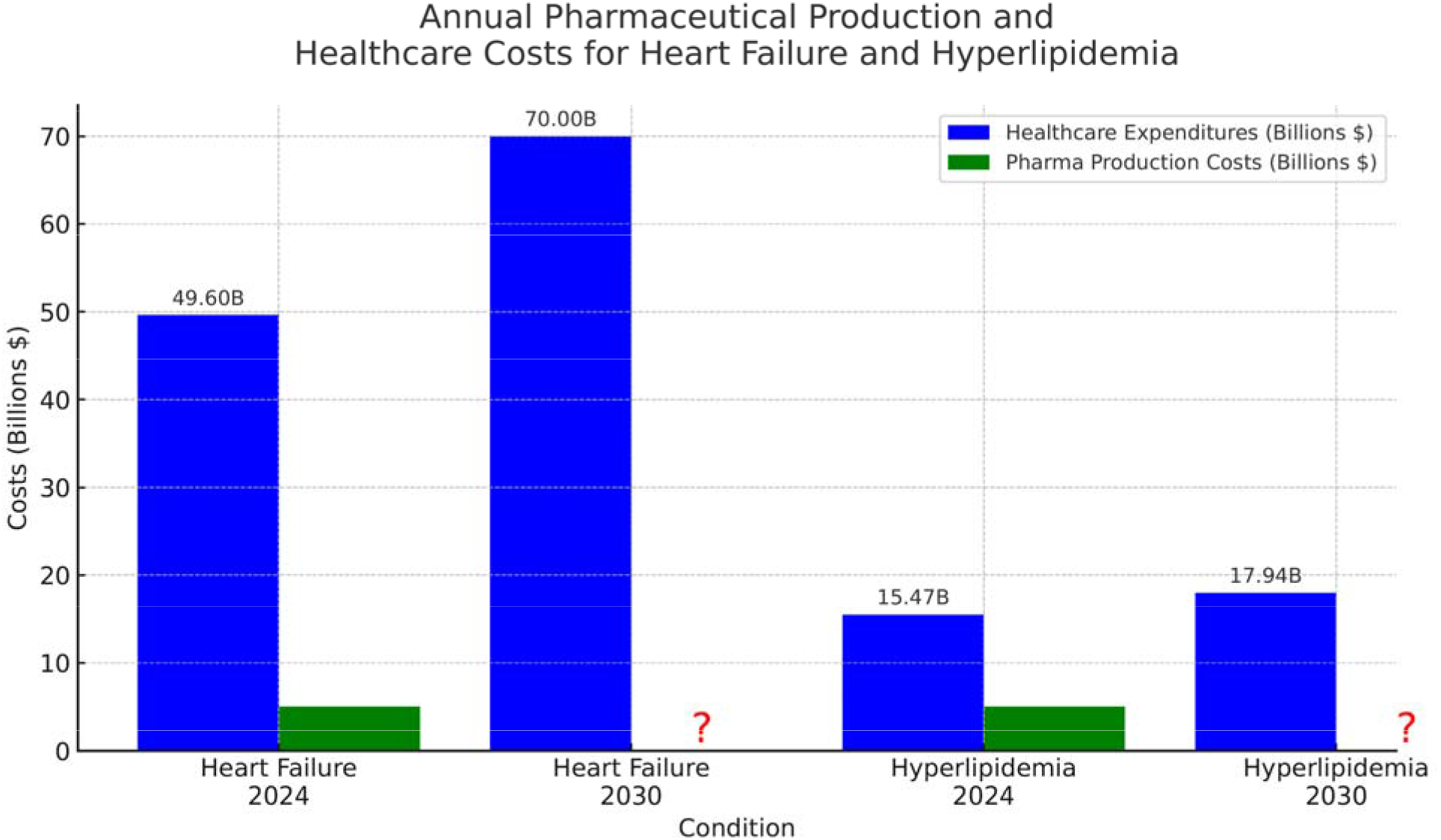
The bar graph illustrates the estimated annual pharmaceutical production costs and healthcare expenditures associated with heart failure and hyperlipidemia for the years 2024 and projections for 2030. For heart failure in 2024, the combined average of estimated direct costs is depicted alongside the projected increase expected by 2030, with pharmaceutical production costs remaining uncertain (denoted by ‘?’). Similarly, for hyperlipidemia, the 2024 expenditures are based on current data, and the 2030 projections are calculated with an assumed annual growth rate of 2.5%, with future pharmaceutical production costs also uncertain.

There is a challenge in bridging the gap between the proven benefits of new drugs and their practical application, especially in private practices constrained by limited resources for continuous education and aggressive marketing strategies of pharmaceutical companies. Additionally, adherence to prescribing guidelines is crucial to ensure the safe and effective use of these medications ^6,7^. The following guidelines help physicians make informed decisions, minimize the risk of adverse effects, and ensure that treatments are used appropriately based on the latest clinical evidence. This adherence is particularly important in private practices, where the pressure of resource limitations and pharmaceutical marketing can lead to deviations from best practices ^8,9^.

Achieving the optimal prescription of these medications involves understanding the diverse approaches of physicians towards new treatments, the role of insurance coverage in facilitating access to costly drugs, and the crucial cooperation of patients with new treatment regimens. Highlighted examples include innovative treatments for heart failure such as sodium-glucose co-transporter-2 (SGLT2) inhibitors, such as empagliflozin ^10^, dapagliflozin ^11^, sotagliflozin ^12^ and cyclic guanosine monophosphate stimulators such as vericiguat ^13^; groundbreaking hyperlipidemia treatments such as evolocumab ^14^, alirocumab ^15^, bempedoic acid ^16^ and inclisiran ^17^ and other cardiac medications such as Mavacampten for hypertrophic cardiomyopathy ^18^ or Rilonacept for recurrent pericarditis ^19^ which are setting new standards in disease management ^14,17,20,21^. These developments are critical for addressing chronic conditions that significantly contribute to healthcare costs, morbidity, and mortality. Despite their potential, the integration of these novel treatments into clinical practice faces some potential resistance owing to possible concerns over efficacy, side effects, and cost, among other factors ^8^.

This study is the first to comprehensively explore these barriers within the cardiologist community by assessing their perspectives, aiming to enhance patient care and outcomes for chronic heart failure and hyperlipidemia, and ultimately reduce healthcare expenditures while improving the quality of life of patients globally.

## Method Study

### Design Methods

#### Study Design and Participants

This descriptive study targeted 150 private practice cardiologists in Las Vegas. Of these, 47 voluntarily participated, and 68% (n=32) completed the survey. The study exclusively focused on private practice cardiologists to explore prescribing behaviors distinct from their academic counterparts.

#### Survey Administration

A structured questionnaire was distributed via Typeform (http://www.typeform.com) between January 1 and February 1, 2024. The survey aimed to identify factors influencing the adoption of newly FDA-approved cardiovascular medications.

#### Data Collection Instrument

The questionnaire was meticulously designed to capture both quantitative and qualitative data through a mix of closed- and open-ended questions. A pilot test was conducted with five randomly selected cardiologists to evaluate the validity and reliability of the instrument. Feedback from the pilot informed revisions to improve clarity and added two questions to enhance the comprehensiveness of the survey.

#### Distribution and Follow-Up

The survey link was disseminated to all cardiologists through email, ensuring a direct and personal approach to inviting participation. A strategic follow-up schedule was implemented to maximize response rates. An initial reminder email was dispatched one week after the original invitation, followed by a second reminder two weeks later. The methodological follow-up was designed to encourage participation.

#### Data Analysis

The average ranking of importance for each barrier or concern about prescribing new FDA-approved medications was calculated by first dividing each respondent’s ranked list of barriers into individual items. Each barrier mentioned by a respondent was assigned a rank based on its position in their list, with the first barrier being the most important, and so on. The calculation steps are as follows.

1. **Assign Ranks:** For respondent, barriers were ranked from most important (rank 1) to least important based on their order of mention. If a respondent listed barriers in a specific sequence, that sequence was used to assign numerical ranks, where a lower number indicated a higher importance.
2. **Sum Ranks for Each Barrier:** The ranks assigned to each barrier across all respondents were summed. This means if “Patient copay amount” was ranked 1 by one respondent and 3 by another, its total rank sum (for these two respondents) would be 4.
3. **Count Occurrences:** The number of times each barrier was mentioned by all respondents was recorded. This gives us the frequency of each barrier being listed, which, in this dataset, was equal across all barriers owing to the survey’s structured response format.
4. **Calculate Average Ranking:** The average ranking of importance for each barrier was calculated by dividing the total rank sum (from Step 2) by the number of occurrences (from Step 3). This gives an average rank for each barrier, where a lower average rank indicates a higher perceived importance.

(Average Ranking=Total Rank Sum for a Barrier/Number of Occurrences of the Barrier)

The average ranking reflects the collective assessment of the barriers’ importance by all respondents, providing a measure that considers both the frequency of mentions and the priority level assigned by those mentioning them.

To categorize the responses, we first identified common themes and then grouped the similar factors into categories. Common categories include the following.

- Economic factors include cost, insurance coverage, affordability, and cost effectiveness.
- Clinical Factors: Covering efficacy, clinical trials, clinical benefits, clinical outcomes, and evidence-based outcomes.
- Safety factors include side effects, safety profiles, adverse effects, and drug-drug interactions.
- Guidelines/recommendations: Encompassing guideline recommendations, supporting data, and evidence-based medicine.
- Patient-centered Factors: Such included affordability, characteristics, compliance, and tolerability.

The collected data were analyzed, and figures were generated using GraphPad software and a custom script in Python, leveraging the Matplotlib library for visualization. This analysis involved descriptive statistics were used to summarize the demographic characteristics of the respondents and their responses to the questionnaire items. Statistical significance was determined using chi-square tests or Fisher’s exact tests, with p-values <0.05 considered significant.

## Results

The study achieved a response rate of 68 %, with 32 of the 47 contacted cardiologists completing the questionnaire. Initial responses increased from 42% after the first reminder to 68% after the third reminder, demonstrating the efficacy of follow-up reminders in boosting participation. The respondent demographics spanned from early career to seasoned professionals, providing a wide range of experience levels.

Our analysis focused on 16 potential barriers, presented in Figure 2, identifying six barriers as top priorities. “Prior authorization requirements” emerged as the most frequently cited top barrier, significantly more than “Medication mechanism of action” and “Patient’s insurance plan” (p=0.01 and p=0.002, respectively) (Figure 2a). “Patient copay amount” and “Overestimation of the benefits of a product” were also prominent, significantly exceeding concerns over “Patient’s insurance plan” (p=0.01) (Figure 2a). The remaining ten barriers were not predominantly chosen as top concerns. In contrast, “Report from leaders in the field” and “Willingness to learn from personal experience” were deemed least important in prescribing new medications. The most significant barriers to prescribing new FDA-approved medications are prior authorization requirements, patient copay amounts, and overestimation of the benefits of a product. Additionally, the expected time for prior authorization, patients’ insurance plans, and medication mechanism of action also played a significant role in influencing prescribing decisions (Figure 2b). Additionally, responses to an open-ended question revealed “Economic Factors” as the dominant consideration over clinical or safety concerns when prescribing new medications (p<.0001) (Figure 2c). This finding underscores the significant impact of cost and insurance coverage on prescription behavior. The analysis indicated that economic considerations significantly outweigh guidelines/recommendations, patient-centered factors, and even clinical efficacy, emphasizing their pivotal role in the decision-making process. The methodology for Figures 2b and 2c is described in detail in the Methods section.

**Figure 2.**
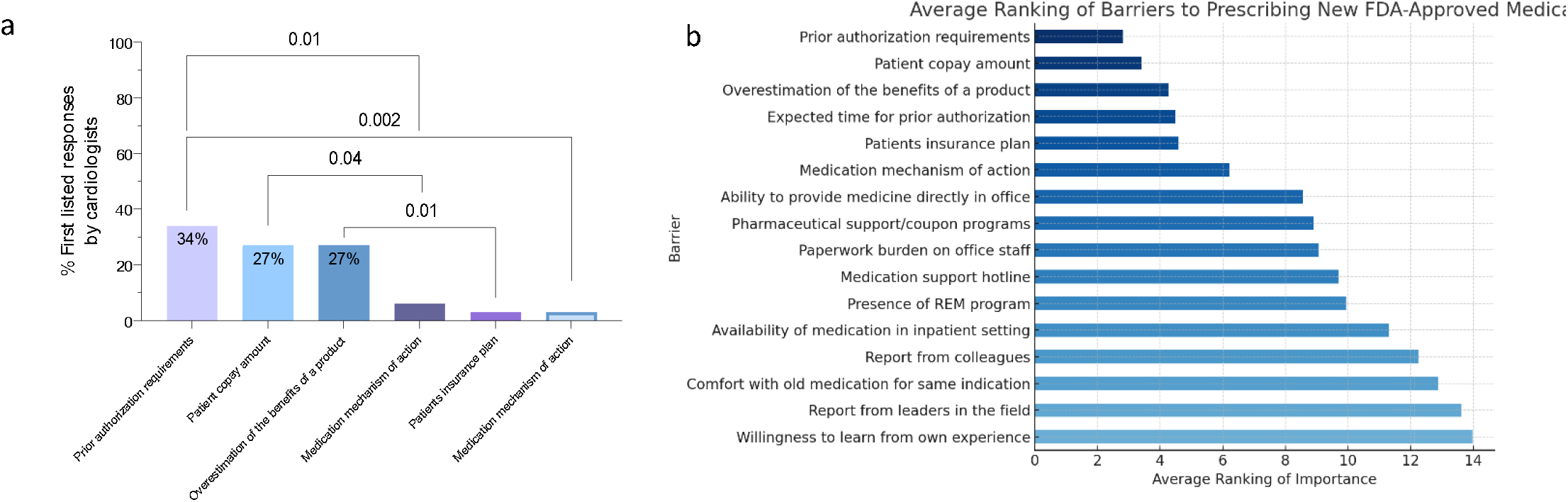
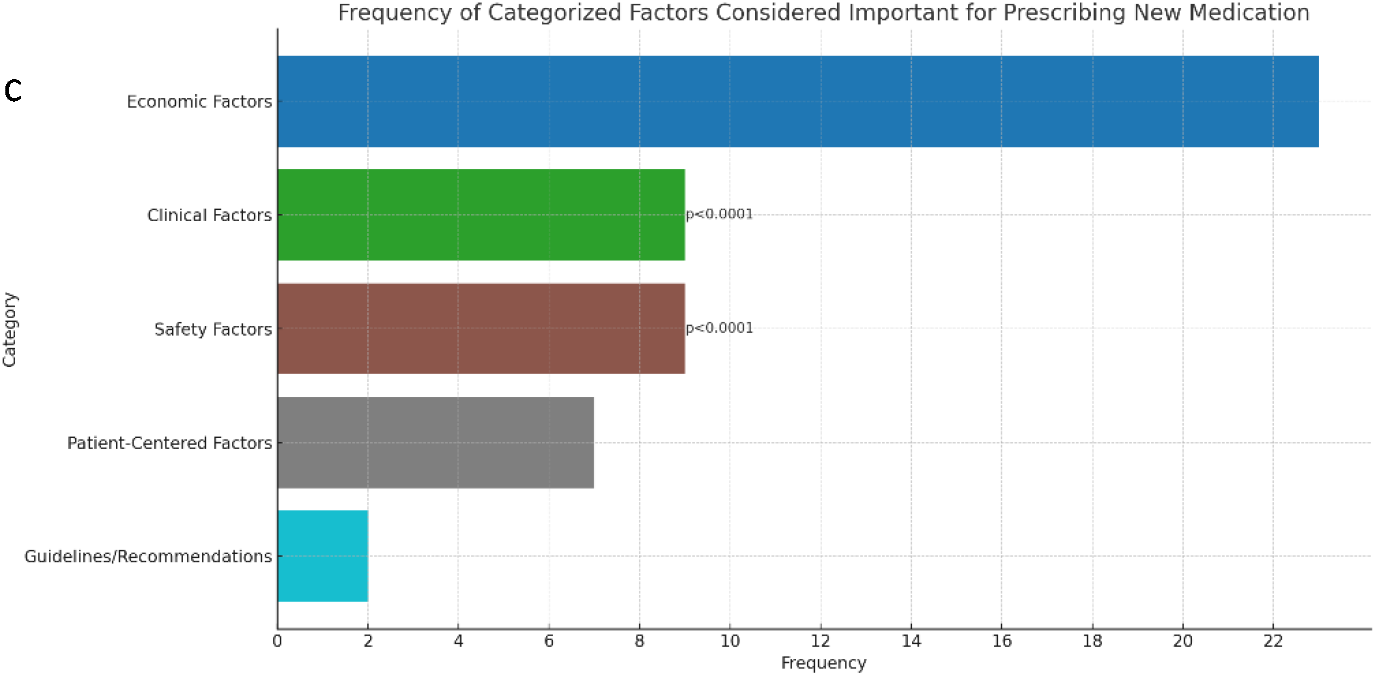
Factors deemed important by cardiologists when deciding to prescribe new FDA-approved medications. **a**. The frequency of first listed answers by cardiologists in percentage. Two-tailed Fisher’s Exact Test was conducted. **b**. The figure is a bar plot showing the barriers to prescribing new FDA-approved medications, ranked by their average importance as indicated by 32 respondents. The x-axis represents the average ranking of importance, with a lower number indicating a higher priority. The y-axis lists the barriers. **c**. The figure is a bar plot that shows the frequency of categorized factors considered important when medical professionals are deciding to prescribe a new medication. The categories, which represent aggregated themes, are listed on the y-axis, and their corresponding frequencies are on the x-axis. This visualization indicates which categories of factors are prioritized b medical professionals in the decision-making process for prescribing new medications. P-values are from comparing with “economic factors”. Two-tailed Fisher’s Exact Test was conducted.

Data from Figure 3a reveals that the majority of cardiologists, 54%, begin prescribing new medications within a “2-6 months” timeframe, a preference that is significantly greater than starting within “1 month,” “6-12 months,” or “2 months” (p=0.01, 0.01, and <0.0001, respectively). About 20% of cardiologists report initiating prescriptions within “1 month” and %54 within the “6-12 months” period, indicating a general tendency to wait at least two months before adopting new medications into their practice. Furthermore, our findings highlight that cardiologists often incorporate new medications into their practice, a behavior significantly more frequent than prescribing “Sometimes” or “Rarely” (p < 0.0001 for both comparisons) as shown in Figure 3b. This indicates a notable readiness among cardiologists to embrace new treatments, suggesting a commitment to leverage the latest clinical advancements and regulatory endorsements to offer cutting-edge care. Additionally, our analysis found no significant difference in the frequency of prescribing new medications among cardiologists with varying practice durations (Figure 3c). This implies that the inclination to prescribe newly approved drugs is broadly consistent across professions, regardless of experience level, and reflects collective confidence in the efficacy and safety of new treatments approved by regulatory authorities.

**Figure 3.**
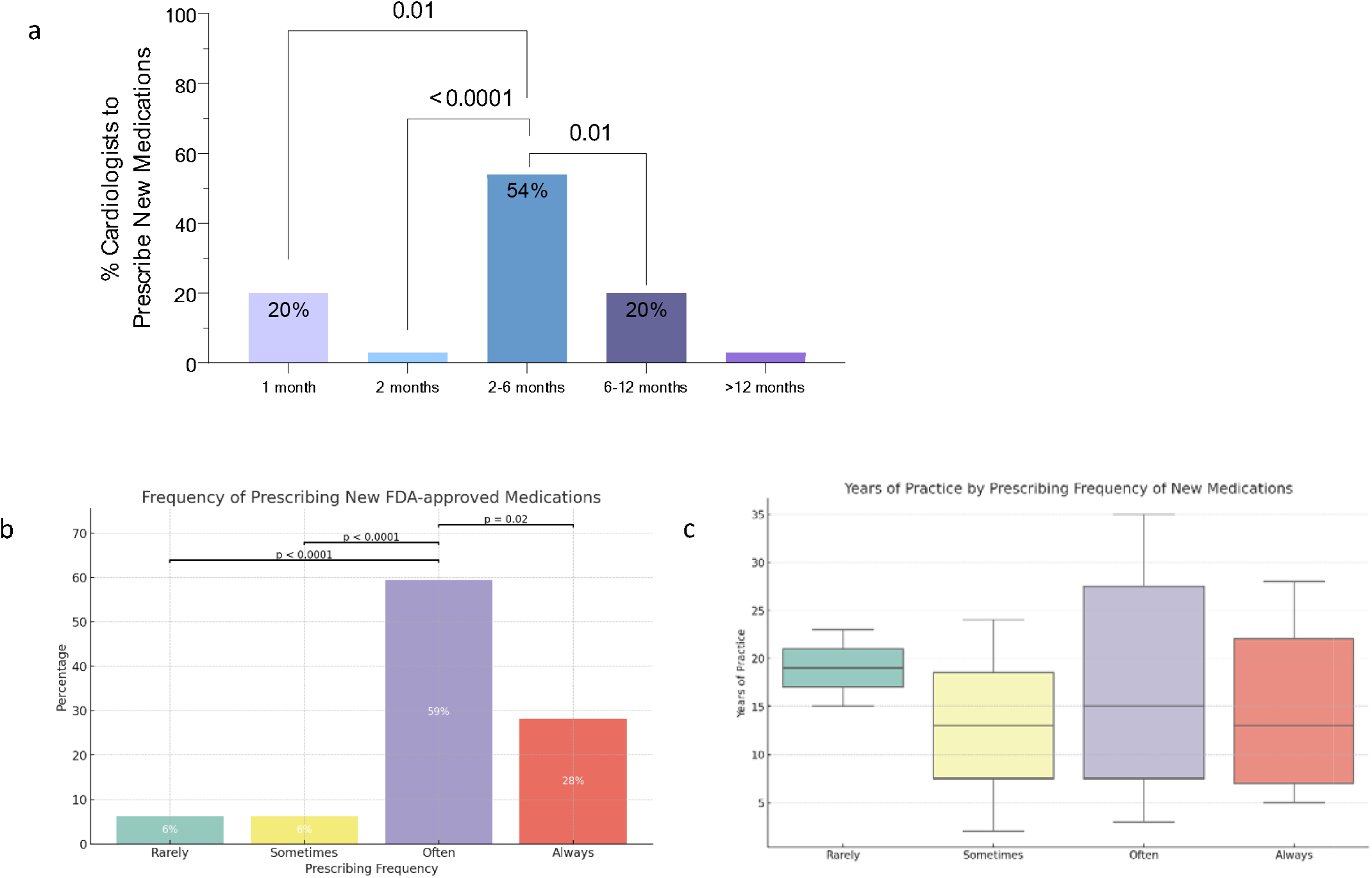
Distribution of cardiologists’ timeframes and frequency for prescribing new FDA-approved medications. **a**. The bar chart illustrates the distribution of the time it takes for cardiologists to begin prescribing new FDA-approved medications. The time intervals are presented on the x-axis, ranging from “1 month” to “>12 months.” The y-axis represents the percentage of cardiologists that correspond to each time interval. Each bar is annotated with the percentage of cardiologists and is color-coded to differentiate between the time intervals. P-values are displayed above brackets spanning the bars to indicate the statistical significance of differences between the adjacent groups. Two-tailed Fisher’s Exact Test was conducted. **b**. The bar chart shows the frequency of prescribing new FDA-approved medications by cardiologists, labeled with “Rarely,” “Sometimes,” “Often,” and “Always.” Each bar’s height represents the count of cardiologists in each category, with whole-number percentages displayed in the center of the bars for clarity. A dashed horizontal line is drawn above the “Often” and “Sometimes” categories, with the p-value from Fisher’s Exact Test displayed above, indicating a statistically significant difference between these two prescribing frequencies. Two-tailed Fisher’s Exact Test was conducted. **c**. The boxplot displays the distribution of years of practice among cardiologists categorized by their frequency of prescribing new FDA-approved medications.

We conducted a descriptive analysis and identified 15 distinct and effective methods through which respondents learned about new medications, with an average preference occurrence of 1.93 (Figure 4a). Preferences varied from one mention to a maximum of six mentions for the most favored method. “Conferences and seminars” emerged as the top choice for staying informed about new drugs, followed by “Medical journals,” “Pharmaceutical representatives,” and “Online webinars.” Further analysis highlighted a clear preference hierarchy, with “Conferences and seminars” being the predominant source of information. This preference is statistically significant when compared to “Pharmaceutical representatives” (p=0.01) and “Online webinars” (p=0.04) (Figure 4), indicating a strong preference for traditional and formal sources of continuing medical education over interactions with the pharmaceutical industry and digital platforms.

**Figure 4.**
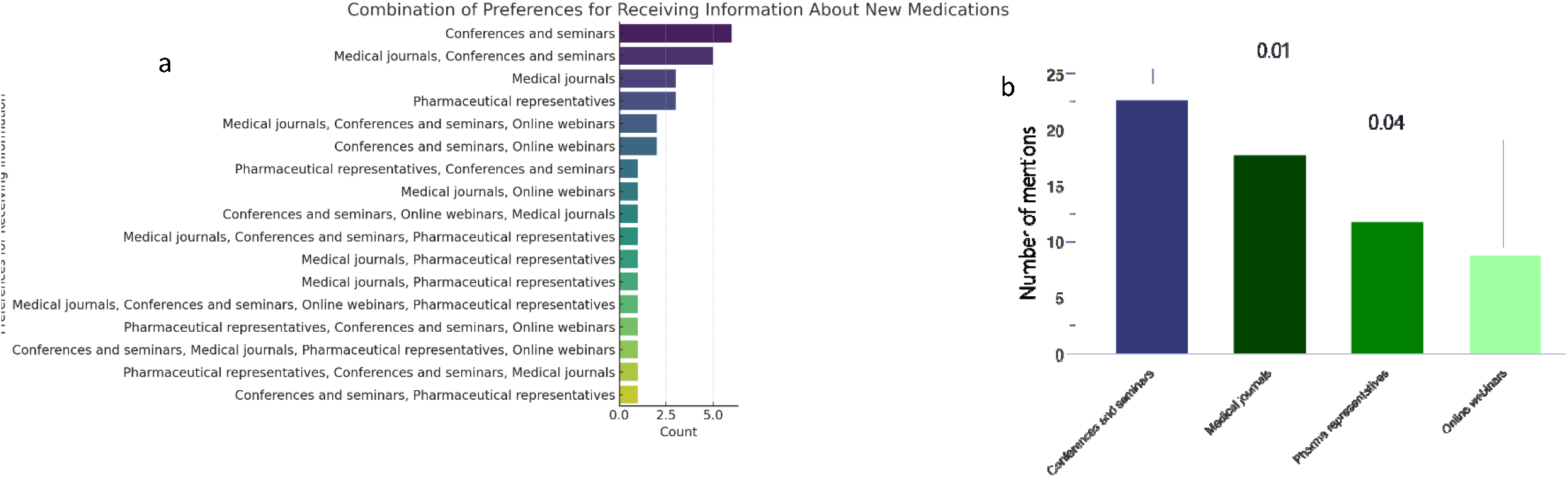
Cardiologists’ preferred methods for receiving information about new medications. **a**. The figure illustrates the distribution of cardiologists’ preferences for receiving information about new medications. The vertical axis lists the various preferred methods, while the horizontal axis shows the count of respondents for each method. The distribution reflects a diverse set of preferences, highlighting the multifaceted approach cardiologists employ to keep abreast of advancements in medical treatment options. **b**. The bar plot visualizes the frequency of mentions for each preference related to receiving information about new medications among respondents. The preferences are listed on the y-axis, and the corresponding number of mentions is on the x-axis. Two-tailed Fisher’s Exact Test was conducted.

## Discussion

Our study highlighted that a significant proportion of cardiologists (54%) preferred to incorporate new FDA-approved medications into their practice within 2-6 months, favoring traditional conferences for learning about these medications. Financial considerations were identified by 61% of respondents as a primary factor influencing their prescription decisions, underscoring the critical role of economic aspects in the adoption of new therapies. Prior authorization, which is a time-consuming and costly process for private practices, significantly impacted prescription habits. Similarly, the surveyed physicians viewed Risk Evaluation and Mitigation Strategy (REMS) programs as deterrents.

Our study highlights the complex and multifactorial nature of the decision-making process in prescribing new medications and identifies economic concerns as a primary factor. Echoing our findings, Medlinskiene et al. (2021) emphasized the multifaceted influences on this process, including patient education, engagement, preferences, medication costs, reimbursement policies, and clinical guidelines ^22^. Their extensive meta-analysis revealed a wide spectrum of factors affecting the adoption of new medications in healthcare settings, pinpointing the cost of new treatments and limited healthcare funding as significant barriers, aligning closely with our observations ^22^.

In contrast, Anderson et al. (2018) found that a substantial majority of physicians (63.8%) did not adopt any new drugs within the initial 15 months of availability, with physician specialty and sex being key determinants of this trend ^23^. However, our research indicates that the majority of cardiologists (54%) are inclined to begin prescribing new medications within a timeframe of 2-6 months, without noting any significant correlation between age and the likelihood of prescribing new treatments. This discrepancy may reflect a shift in physician satisfaction with the development of new drugs over time, or it might be attributable to our specific focus on the perspectives of cardiologists, who seem predisposed to quicker adoption of innovative treatments, or possibly because of our study’s concentration on a select group of private practice cardiologists in a localized area. Bourke and Roper’s (2012) study in Ireland highlighted slower drug adoption rates among older GPs and those without support staff, suggesting targeted support for these groups ^24^. However, our findings did not focus on any correlation between cardiologists’ age and their prescribing frequency.

Favoritism towards conferences and seminars likely mirrors a wider appreciation within the medical field for peer interaction and the authoritative value of professional events. This preference is likely motivated by the depth of learning, discussion, and critical analysis offered by these forums, which can significantly impact clinical practice. Interestingly, we noted a lower preference for pharmaceutical representatives, possibly reflecting a cautious stance towards information provided by the industry or an effort to mitigate bias in decision-making, as evidenced by the identification of overestimation of a product’s benefits as a barrier in Figure 2. The notable discrepancy in the preference for conferences over interactions with pharmaceutical representatives indicates that while the latter are considered less desirable, they maintain a role in communicating information about new drugs. The minimal preference for online webinars may stem from their perceived lack of interactive and networking opportunities, which are highly valued in traditional gatherings. Nonetheless, the shift towards digital platforms in medical education, hastened by events such as the COVID-19 pandemic, hints at potential changes in these preferences over time. The diversity of favored sources suggests that cardiologists employ a varied array of channels to stay updated on new medications, advocating for a comprehensive strategy to assimilate and apply new medical insights.

A key strength of our study was its innovative approach, utilizing a questionnaire to gather cardiologists’ perspectives, in contrast to studies such as that of Anderson et al. (2018), who analyzed monthly prescribing data across all practicing physicians ^23^. The inclusion of both open- and closed-ended questions provided consistent results, serving as an internal validation of our findings. Furthermore, the use of a cloud-based platform for data collection and analysis enhanced the integrity of the data and facilitated respondent participation. Notably, our study achieved a high response rate in a short period of time, which is uncommon in survey-based research. This, along with the varied experience levels of the cardiologists who participated, strengthens the generalizability and reliability of our conclusions. Additionally, we not only explored barriers to prescribing new medications, but also sought cardiologists’ suggestions on enhancing their willingness to adopt new treatments, offering practical insights for overcoming these obstacles.

However, our study had several limitations. Notably, the sample size and homogeneity of the surveyed population were limiting factors. Future research should address these issues by including a larger, more diverse cohort across different geographical locations. As the study was conducted in Las Vegas, the findings may not accurately reflect the overall prescribing practices of cardiologists across the United States. Unlike other studies, we did not categorize responses based on different classes of drugs, which could have revealed specific trends in prescribing behaviors, particularly regarding newly approved drugs, for both new and old indications.

Another limitation is the reliance on self-reported behaviors, which raises concerns about the alignment of these reports with actual prescribing practices. Additionally, patient characteristics that influence prescription decisions have not been deeply explored. For instance, Min Zhao et al. (2020) highlighted sex differences in the prescription of cardiovascular medications, underscoring the importance of evaluating how such factors shape prescribing habits ^25^. Addressing these aspects in future research will be critical for a more comprehensive understanding of the dynamics at play in physicians’ prescribing behaviors. To gain a more comprehensive understanding of the factors influencing medication adoption, future research could benefit from incorporating patient surveys. This would provide valuable insights into patients’ perspectives on medication cost, access, and adherence, complementing the data collected from cardiologists.

In conclusion, our study reflects the eagerness of cardiologists to adopt new medications promptly, driven by the potential to improve patient outcomes and reduce healthcare costs. However, overcoming financial barriers in the prescribing process remains a crucial challenge. Future research should focus on longitudinal studies to track changes in prescribing behaviors in response to healthcare policy and insurance coverage updates and the role of digital platforms in shaping medical education and information dissemination.

## Conclusion

In conclusion, our research underscores a proactive stance among cardiologists toward the integration of new cardiac medications into clinical practice, reflecting a positive outlook on embracing changes and advancements in treatment options. Despite enthusiasm to adopt cutting-edge therapies, the decision-making process is heavily influenced by economic considerations, highlighting the significant role of patient affordability and third-party payer policies in the practical application of new guidelines. A delicate balance between clinical innovation and economic feasibility is needed for strategic approaches to ensure that advancements in cardiac care are accessible to those in need, ultimately aiming to improve patient outcomes within the constraints of the economic realities of the healthcare system.

## Data Availability

All data produced in the present study are available upon reasonable request to the authors

## Availability of data and material

The data supporting this study will be made available upon reasonable request.

## Code availability

Not applicable.

## Authors’ contributions

L.V. was responsible for conceptualizing the study and conducting the first and second reviews. A.M. served as the project leader, overseeing its development from its inception, managing all aspects of the project, and contributing the most substantial efforts. A.Mo. participated in the data analysis and drafting of the manuscript. S.G. contributed to the preparation and refinement of the first and second drafts of this manuscript.

## Consent to participate

Not applicable.

## Consent for publication

Not applicable.

## Ethical Considerations and Institutional Review Board Waiver

Identifiable information will be anonymized to protect the cardiologists’ privacy. Given the non-invasive nature of this research, an Institutional Review Board (IRB) waiver will be sought. The waiver request argues that the study does not involve vulnerable populations, medical interventions, or access to participants’ personal records beyond what is necessary for the research objectives.

## Reference

1 Tamargo, J. et al. New pharmacological agents and novel cardiovascular pharmacotherapy strategies in 2022. Eur Heart J Cardiovasc Pharmacother 9, 353–370, doi:10.1093/ehjcvp/pvad034 (2023).

2 Heidenreich, P. A. et al. Economic Issues in Heart Failure in the United States. J Card Fail 28, 453–466, doi:10.1016/j.cardfail.2021.12.017 (2022).

3 Zhang, D., Wang, G., Fang, J. & Mercado, C. Hyperlipidemia and Medical Expenditures by Cardiovascular Disease Status in US Adults. Med Care 55, 4–11, doi:10.1097/mlr.0000000000000663 (2017).

4 Jafari, L. A., Suen, R. M. & Khan, S. S. Refocusing on the Primary Prevention of Heart Failure. Curr Treat Options Cardiovasc Med 22, doi:10.1007/s11936-020-00811-3 (2020).

5 Zhang, D., Wang, G., Fang, J. & Mercado, C. Hyperlipidemia and Medical Expenditures by Cardiovascular Disease Status in US Adults. Medical Care 55, 4–11, doi:10.1097/mlr.0000000000000663 (2017).

6 Mohammadi, A. et al. Abstract 4142032: Hospital Discharge Education For Heart Failure Patients Delivered by Conversational Agent Technology Improves Patient Experience and Is Preferred to a Doctor or Nurse. Circulation 150, A4142032-A4142032, doi:doi:10.1161/circ.150.suppl_1.4142032 (2024).

7 Mosca, L. et al. Effectiveness-based guidelines for the prevention of cardiovascular disease in women--2011 update: a guideline from the american heart association. Circulation 123, 1243–1262, doi:10.1161/CIR.0b013e31820faaf8 (2011).

8 Haeussler, C. & Assmus, A. Bridging the gap between invention and innovation: Increasing success rates in publicly and industry-funded clinical trials. Research Policy 50, 104155, doi:10.1016/j.respol.2020.104155 (2021).

9 Masoompour, S. M., Mohammadi, A. & Mahdaviazad, H. Adherence to the Global Initiative for Chronic Obstructive Lung Disease guidelines for management of COPD: a hospital-base study. The Clinical Respiratory Journal 10, 298–302, doi:10.1111/crj.12215 (2016).

10 Anker, S. D. et al. Empagliflozin in Heart Failure with a Preserved Ejection Fraction. New England Journal of Medicine 385, 1451-1461, doi:doi:10.1056/NEJMoa2107038 (2021).

11 McMurray, J. J. V. et al. Dapagliflozin in Patients with Heart Failure and Reduced Ejection Fraction. New England Journal of Medicine 381, 1995-2008, doi:doi:10.1056/NEJMoa1911303 (2019).

12 Bhatt, D. L. et al. Sotagliflozin in Patients with Diabetes and Recent Worsening Heart Failure. New England Journal of Medicine 384, 117-128, doi:doi:10.1056/NEJMoa2030183 (2021).

13 Armstrong, P. W. et al. Vericiguat in Patients with Heart Failure and Reduced Ejection Fraction. New England Journal of Medicine 382, 1883-1893, doi:doi:10.1056/NEJMoa1915928 (2020).

14 Sabatine, M. S. et al. Evolocumab and Clinical Outcomes in Patients with Cardiovascular Disease. New England Journal of Medicine 376, 1713-1722, doi:doi:10.1056/NEJMoa1615664 (2017).

15 Schwartz, G. G. et al. Alirocumab and Cardiovascular Outcomes after Acute Coronary Syndrome. New England Journal of Medicine 379, 2097-2107, doi:doi:10.1056/NEJMoa1801174 (2018).

16 Ray, K. K. et al. Efficacy and safety of bempedoic acid among patients with and without diabetes: prespecified analysis of the CLEAR Outcomes randomised trial. Lancet Diabetes Endocrinol 12, 19–28, doi:10.1016/s2213-8587(23)00316-9 (2024).

17 Ray, K. K. et al. Two Phase 3 Trials of Inclisiran in Patients with Elevated LDL Cholesterol. New England Journal of Medicine 382, 1507-1519, doi:doi:10.1056/NEJMoa1912387 (2020).

18 Spertus, J. A. et al. Mavacamten for treatment of symptomatic obstructive hypertrophic cardiomyopathy (EXPLORER-HCM): health status analysis of a randomised, double-blind, placebo-controlled, phase 3 trial. Lancet 397, 2467–2475, doi:10.1016/s0140-6736(21)00763-7 (2021).

19 Klein, A. L. et al. Phase 3 Trial of Interleukin-1 Trap Rilonacept in Recurrent Pericarditis. New England Journal of Medicine 384, 31-41, doi:doi:10.1056/NEJMoa2027892 (2021).

20 Solomon, S. D. et al. Dapagliflozin in Heart Failure with Mildly Reduced or Preserved Ejection Fraction. New England Journal of Medicine 387, 1089-1098, doi:doi:10.1056/NEJMoa2206286 (2022).

21 Gheorghiade, M. et al. Effect of Vericiguat, a Soluble Guanylate Cyclase Stimulator, on Natriuretic Peptide Levels in Patients With Worsening Chronic Heart Failure and Reduced Ejection Fraction: The SOCRATES-REDUCED Randomized Trial. JAMA 314, 2251–2262, doi:10.1001/jama.2015.15734 (2015).

22 Medlinskiene, K. et al. Barriers and facilitators to the uptake of new medicines into clinical practice: a systematic review. BMC Health Serv Res 21, 1198, doi:10.1186/s12913-021-07196-4 (2021).

23 Anderson, T. S. et al. Patterns and predictors of physician adoption of new cardiovascular drugs. Healthc (Amst) 6, 33–40, doi:10.1016/j.hjdsi.2017.09.004 (2018).

24 Bourke, J. & Roper, S. In with the new: the determinants of prescribing innovation by general practitioners in Ireland. Eur J Health Econ 13, 393–407, doi:10.1007/s10198-011-0311-5 (2012).

25 Zhao, M. et al. Sex Differences in Cardiovascular Medication Prescription in Primary Care: A Systematic Review and Meta-Analysis. J Am Heart Assoc 9, e014742, doi:10.1161/jaha.119.014742 (2020).

